# Pulmonary and Cardiac Pathology in Covid-19: The First Autopsy Series from New Orleans

**DOI:** 10.1101/2020.04.06.20050575

**Authors:** Sharon E. Fox, Aibek Akmatbekov, Jack L. Harbert, Guang Li, J. Quincy Brown, Richard S. Vander Heide

**Affiliations:** Department of Pathology, LSU Health Sciences Center, New Orleans; Pathology and Laboratory Medicine Service, Southeast Louisiana Veterans Healthcare System; Department of Biomedical Engineering, Tulane University

## Abstract

SARS-CoV-2 has rapidly spread across the United States, causing extensive morbidity and mortality, though the histopathologic basis of severe disease cases has yet to be studied in detail. Over the past century, autopsy has contributed significantly to our understanding of numerous disease processes, but for several reasons, autopsy reports following deaths related to SARS-CoV-2 have thus far been limited across the globe. We report on the relevant cardiopulmonary findings in the first series of autopsies in the United States, with the cause of death being due to SARS-CoV-2 infection. These cases identify key pathologic states potentially contributing to severe disease and decompensation in these patients.

## Introduction

The first confirmed case of SARS-CoV-2 infection in the United States was reported on January 20, 2020. Since that time, the virus has spread across the country, with several cities within the United States becoming epicenters of the pandemic. As of March 31, 2020 the Louisiana Department of Health reported a total of 5,237 COVID-19 cases with 1,355 hospitalizations, and 239 COVID-19 related deaths statewide. A total of 1,834 of the 5,239 COVID-19 cases and 101 of the 239 deaths have occurred in the city of New Orleans – the highest rate of death per capita in the United States. University Medical Center in New Orleans, built following Hurricane Katrina, is equipped with an autopsy suite meeting the modern standards recommended by the CDC for performance of autopsy on COVID-19 positive patients. We report here on the cardiopulmonary findings of the first four autopsies of a series of twelve performed on patients within the United States, with relevant implications for the treatment of severe cases.

## Brief Clinical Summary

The four decedents included male and female patients, ages 44-76. All were African American, and had a history of obesity class 2-3, and hypertension controlled by medication. Three of the patients had insulin-dependent type II diabetes, two had known chronic kidney disease (stages 2 and 3), and one was taking methotrexate.

In all cases the clinical course consisted of approximately three days of mild cough and fever to 101- 102° F., with sudden respiratory decompensation just prior to arrival in the emergency department. Chest radiographs revealed bilateral ground-glass opacities, consistent with acute respiratory distress syndrome (ARDS) which worsened over the hospital course. The patients were intubated and brought to the ICU. Treatment in the ICU included vancomycin, azithromycin, and aefepime for all patients, with one patient receiving dexamethasone. All of the patients tested positive for SARS-CoV-2 (by 2019 Novel Coronavirus Real Time RT-PCR).

Notable laboratory findings were the development of elevated ferritin, fibrinogen, PT, and within 24 hours of death, an increased neutrophil count with relative lymphopenia. Glucose and AST became slightly elevated above normal, and creatinine increased above baseline for all patients. D-dimers drawn near the time of death in two patients were markedly elevated (1200-2900 ng/mL). (A detailed description of ante-mortem laboratory findings can be found in **Table S1** in the Supplementary Appendix). When the patients continued to deteriorate despite support, the families elected to withdraw care. In each case, consent for autopsy was given, and non-restricted by the next of kin. Studies performed outside of routine autopsy were determined to be exempt by the IRB at Tulane University.

## Gross Findings

Gross examination of the lungs at the time of autopsy revealed the tracheae to be of normal caliber and mildly erythematous. All of the lungs were heavy, the left ranging from 680g to 1030g, normal (583 +/-216); right ranging from 800g to 1050g, normal (663+/-239). They contained the usual lobes and fissures, with exception to one decedent with prior partial lobectomy on the right side. The pulmonary arteries at the hilum of each of the lungs were free of thromboemboli. The bronchi revealed thick, white mucous in the lungs of one patient, and pink froth in the airways of the other three. Mild to moderate serosanguinous pericardial and pleural effusions were also present. The parenchyma of each of the lungs was diffusely edematous and firm, consistent with the clinical diagnosis of ARDS. Notably, regions of dark-colored hemorrhage with focal demarcation could be identified throughout the peripheral parenchyma in the lungs of all but one of the decedents (**Figure 1A**). On cut sections, the areas identified as hemorrhagic on the external surface showed frank hemorrhage. After fixation, the cut surfaces of the lung tissue showed alternating areas of tan-grey consolidation with patchy areas of hemorrhage that ranged from 3-6 cm in maximal diameter. In some cases, small, firm thrombi were present in sections of the peripheral parenchyma (**Figure 1C**). Only in the case of the patient on immunosuppression was there focal consolidation - the remainder of the lungs showed no evidence of lobar infiltrate, abscess, or definitive gross inflammatory process.

**FIGURE 1:**
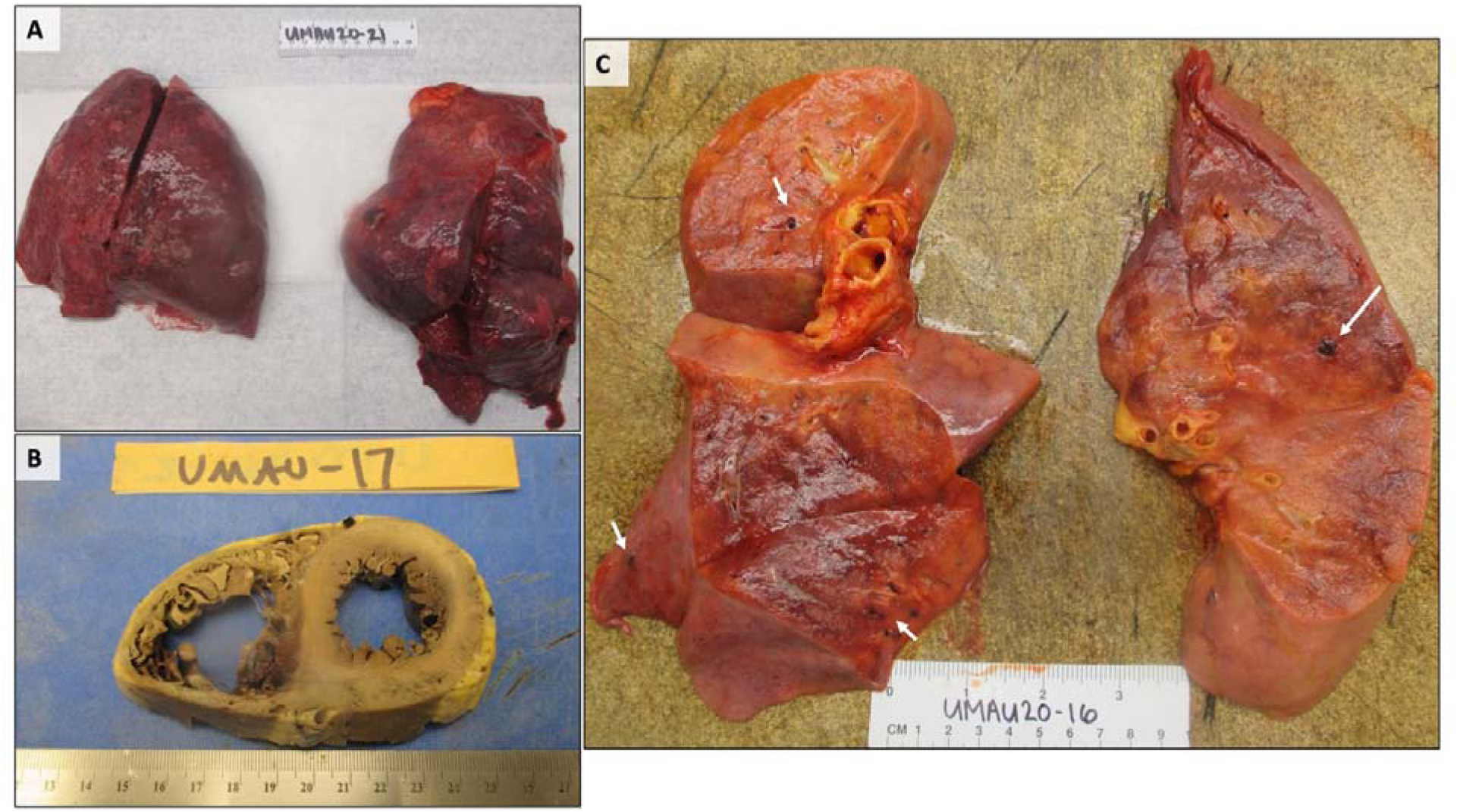
Gross Findings of the Lungs and Heart. **A)** Lungs with bilateral pulmonary edema and patches of dark hemorrhage, and **B)** A heart showing extreme right ventricular dilatation, with straightening of the interventricular septum. **C)** Cut sections of lung showing thrombi present within peripheral small vessels (white arrows).

Examination of the heart was performed in three cases, with the hearts ranging in size from 430g to 550g (normal: 365g +/-71). The most significant gross findings were cardiomegaly, and right ventricular dilatation. In one case, massive dilatation could be seen, in which the right ventricular cavity measured 3.6cm in diameter, while the left ventricle measured 3.4cm in greatest diameter (**Figure 1B**). The cut surface of the myocardium was firm, red-brown, and free of significant lesions in all cases, and the coronary arteries showed no significant stenosis or acute thrombus formation.

## Microscopic Findings

### Pulmonary

The lungs were extensively sampled across central and peripheral regions of each lobe bilaterally. Histologic examination of the lungs showed bilateral diffuse alveolar damage with a comparatively mild-to-moderate lymphocytic infiltrate, composed of a mixture of CD4+ and CD8+ lymphocytes (**Figure 2**), located predominantly in the interstitial spaces and around larger bronchioles. CD4+ lymphocytes could be seen in aggregates around small vessels, some of which appeared to contain platelets and small thrombi. In all but one case, foci of hemorrhage were present. Desquamated type 2-pneumocytes with apparent viral cytopathic effect consisting of cytomegaly, and enlarged nuclei with bright, eosinophilic nucleoli, were present within alveolar spaces (**Figure 3**). The largest of these cells frequently contained an eccentric clearing of the cytoplasm with small vesicles discernible at higher power, likely representing viral inclusions. Scattered hyaline membranes could be seen, as well as fibrin deposition, highlighted by trichrome stains (**Figure 2**), consistent with diffuse alveolar damage. The alveolar capillaries were notably thickened, with surrounding edema, and fibrin thrombi were present within the capillaries and small vessels. A notable finding was the presence of CD61+ megakaryocytes (**Figure 2**), possibly representing resident pulmonary megakaryocytes, with significant nuclear hyperchromasia and atypia. These cells were located within alveolar capillaries, and could be seen in association with, and actively producing platelets (**Figure 2**). The fibrin and platelets present within small vessels also appeared to aggregate inflammatory cells, with entrapment of numerous neutrophils. Only in the case of the patient on immunosuppression was there evidence of a focal acute inflammatory infiltrate possibly consistent with secondary infection. The neutrophils in this case, however, were partially degenerated and entrapped in fibers, possibly representing neutrophil extracellular traps (**Figure 3**),^1,2^ and were present in association with clusters of CD4+ mononuclear cells. No significant neutrophilic infiltrate was identified within airways or the interstitium to suggest secondary infection in other cases.

**FIGURE 2:**
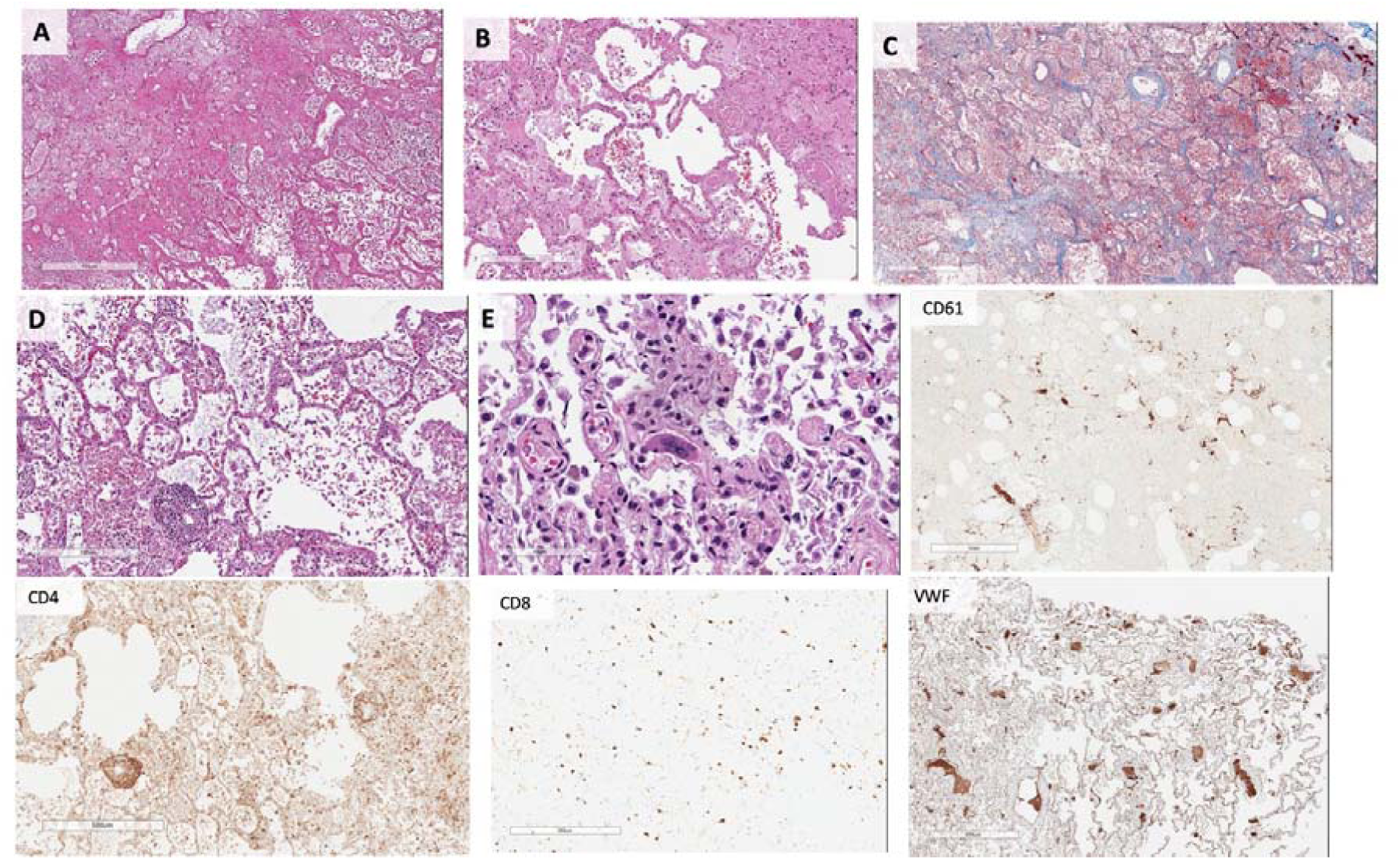
Pulmonary Microscopic Findings. All patients demonstrated extensive diffuse alveolar damage. **A)** Hyaline membranes and hemorrhage (H&E), with **B)** Fibrin thrombi present within distended small vessels and capillaries, and **C)** Extensive extracellular fibrin deposition highlighted in blue by Masson-Trichrome stain. **D)** Perivascular aggregations of lymphocytes, which were positive for CD4 immunostain, with only scattered CD8 positive cells present. **E)** Numerous megakaryocytes were present within the small vessels and alveolar capillaries, highlighted by CD61 and Von Willebrand Factor immunostains.

**FIGURE 3:**
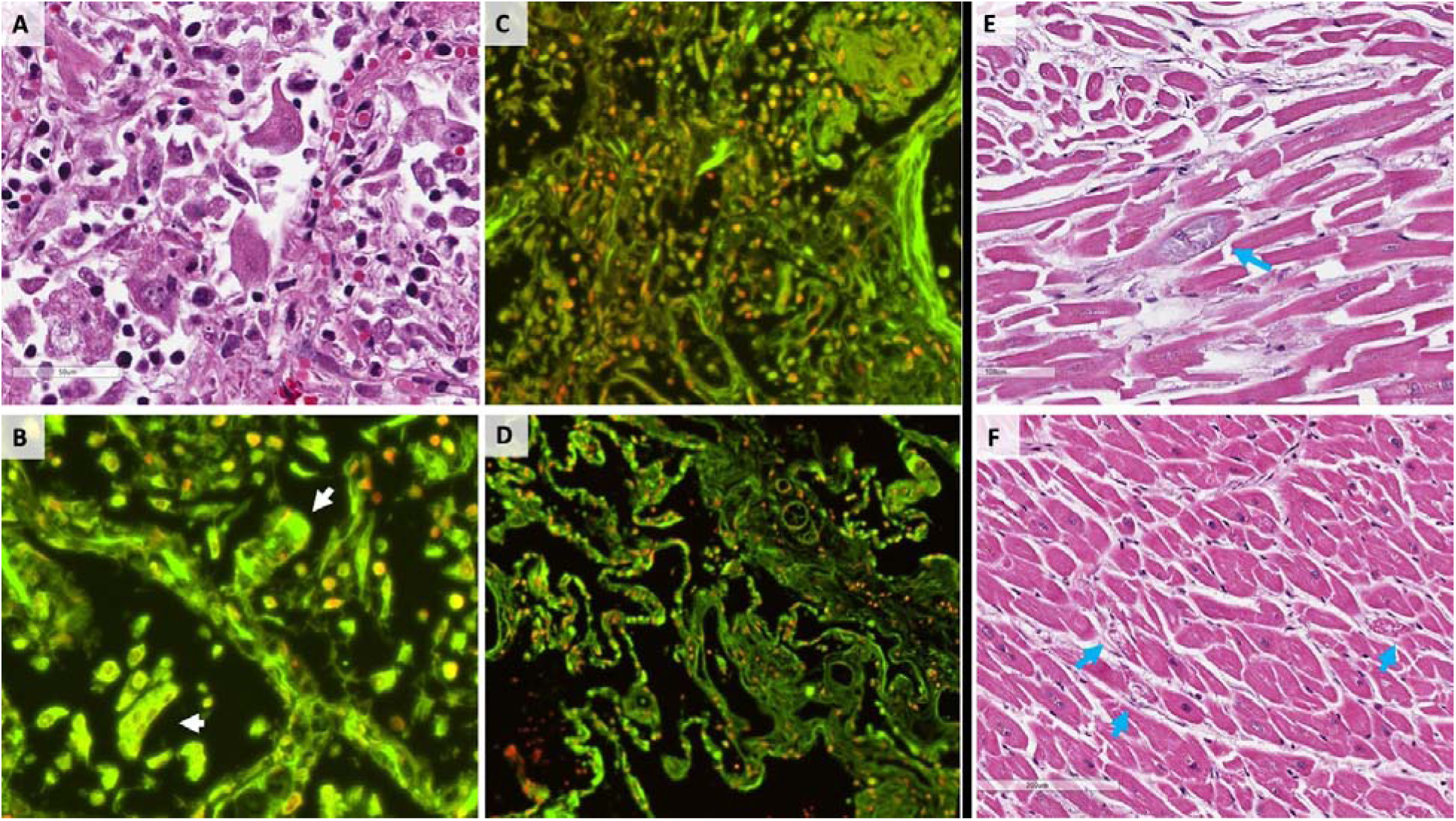
SARS-CoV-2 cytopathic effects. **A)** H&E stain of several enlarged pneumocytes within a damaged alveolus, having enlarged nuclei, prominent nuceloli, and cytologic atypia. **B)** Relative distribution of dsDNA (red) versus RNA (green) in tissue sections via DRAQ5 and SYTO RNASelect fluorescent staining (see **Supplementary Methods** for staining details). Virally infected cells in alveolar spaces show multinucleation and grouping as evidenced by DNA stain, and abundant RNA present within the cytoplasm (white arrows), **C)** Entrapment of immune cells, including degeneration neutrophils, within fibrin, and strands of extracellular material with weak DNA staining, and **D)** Control lung tissue obtained at autopsy for non-pulmonary cause of death prior to the SARS-CoV-2 pandemic. **E)** and **F)** H&E stains of cardiac myocytes with focal degeneration (blue arrows).

### Cardiac

The sections of myocardium did not show any large or confluent areas of myocyte necrosis. The cardiac histopathology was remarkable, however, for scattered individual cell myocyte necrosis in each heart examined. In rare areas, lymphocytes were adjacent to, but not surrounding degenerating myocytes. Whether this may represent an early manifestation of a viral myocarditis is not certain, but there was no significant brisk lymphocytic inflammatory infiltrate consistent with the typical pattern of viral myocarditis. This may be consistent with a recent paper by Chen et al. that hypothesizes that pericytes may be infected by the SARS-CoV-2 virus and cause capillary endothelial cell/microvascular dysfunction which may cause individual cell necrosis.^3^ There was no obvious viral cytopathic effect by light microscopy, but direct viral infection of myocytes cannot be entirely ruled out in this limited examination.

## Discussion

The dominant process in all cases was consistent with diffuse alveolar damage, with a mild to moderate mononuclear response consisting of notable CD4+ aggregates around thrombosed small vessels, and significant associated hemorrhage. Important additional mechanisms that may have contributed to death in this initial series of autopsies include a thrombotic microangiopathy that was restricted to the lungs. This process may involve activation of megakaryocytes, possibly those native to the lung, with platelet aggregation and platelet-rich clot formation, in addition to fibrin deposition. Small vessel thrombus formation in the lung periphery was in many cases associated with foci of alveolar hemorrhage. In one case, extensive fibrin and early organization was present, with degenerated neutrophils within the alveoli possibly representing neutrophil extracellular traps.^1,2^ On RNA imaging, we were able to visualize multinucleated cells within alveolar spaces, containing abundant RNA, likely representing virally infected cells. These may represent the multinucleated cells previously described from a single report of post-mortem biopsy from a decedent in China.^4^ Cardiac findings were significant for a lack of myocarditis, and the rise in BNP observed in at least one of our cases was likely due to acute right ventricular dilatation. The underlying cause of scattered atypical myocyte degeneration remains uncertain.

There is prior evidence of viral infection causing activation of both maladaptive cytokine pathways, and platelet response, and our findings suggest that these immune functions may be related to severe forms of Covid-19. In response to systemic and pulmonary viral infections of H1N1 influenza and dengue, megakaryocytes have been known to respond by overexpressing IFITM3, and producing platelets with the same over-expression ^5^ In addition, platelets and megakaryocytes may have receptors for viruses^6–9^, some of which have been specifically activated in H1N1 influenza, often in association with lymphopenia^10–12^ There is even some evidence that the earlier SARS-CoV directly infected megakaryocytes, and that platelets function was affected in damaged lungs of those with severe SARS.^14^ We do not currently have evidence of direct infection of megakaryocytes by SARS-CoV-2, but the abundance of these cells in the lungs at autopsy is likely related to the abundance of small, sometimes platelet-rich thrombi, and foci of hemorrhage.

A notable finding was the lack of significant secondary infection in all of our cases. While all of the patients received antibiotic therapy throughout their hospital courses, the lack of significant bacterial or fungal infection suggests that this is not the main cause of their decline. We also note that two of our patients were younger than those commonly thought to be ask risk for death due to Covid-19, and without immunosuppressive therapy, though with obesity, hypertension, and diabetes - comorbidities often present in our patient population, and in the population of many cities with Covid-19 on the rise. Based on our findings, we believe that effective therapy for these patients should not only target the viral pathogen, but also the thrombotic and microangiopathic effects of the virus, and possibly a maladaptive immune response to viral infection.

## Data Availability

Data presented within this manuscript is available upon request within applicable law.

## Acknowledgements

We would first like to acknowledge our patients and their families, who in a time of loss have sought to help others understand this disease. We are also grateful for the support of the Department of Pathology at LSU Health Sciences Center, and for the hard work of the University Medical Center staff - in particular, Nicole Bichsel, for her invaluable role as autopsy assistant.

Finally, we would like to acknowledge Dr. Paula L. Bockenstedt, Associate Professor of Hematology at the University of Michigan, for her expertise and guidance in this work.

## References

1. Mikacenic C, Moore R, Dmyterko V, et al. Neutrophil extracellular traps (NETs) are increased in the alveolar spaces of patients with ventilator-associated pneumonia. Crit Care 2018;22(1):358.

2. Lefrançais E, Mallavia B, Zhuo H, Calfee CS, Looney MR. Maladaptive role of neutrophil extracellular traps in pathogen-induced lung injury. JCI Insight [Internet] 2018;3(3). Available from: http://dx.doi.org/10.1172/jci.insight.98178

3. Chen L, Li X, Chen M, Feng Y, Xiong C. The ACE2 expression in human heart indicates new potential mechanism of heart injury among patients infected with SARS-CoV-2. Cardiovasc Res [Internet] 2020;Available from: http://dx.doi.org/10.1093/cvr/cvaa078

4. Xu Z, Shi L, Wang Y, et al. Pathological findings of COVID-19 associated with acute respiratory distress syndrome. Lancet Respir Med [Internet] 2020;Available from: http://dx.doi.org/10.1016/S2213-2600(20)30076-X

5. Campbell RA, Schwertz H, Hottz ED, et al. Human megakaryocytes possess intrinsic antiviral immunity through regulated induction of IFITM3. Blood 2019;133(19):2013–26.

6. Youssefian T, Drouin A, Massé J-M, Guichard J, Cramer EM. Host defense role of platelets: engulfment of HIV and Staphylococcus aureus occurs in a specific subcellular compartment and is enhanced by platelet activation. Blood 2002;99(11):4021–9.

7. Boukour S, Massé J-M, Bénit L, Dubart-Kupperschmitt A, Cramer EM. Lentivirus degradation and DCSIGN expression by human platelets and megakaryocytes. J Thromb Haemost 2006;4(2):426–35.

8. Loria GD, Romagnoli PA, Moseley NB, Rucavado A, Altman JD. Platelets support a protective immune response to LCMV by preventing splenic necrosis. Blood 2013;121(6):940–50.

9. Middleton EA, Weyrich AS, Zimmerman GA. Platelets in Pulmonary Immune Responses and Inflammatory Lung Diseases. Physiol Rev 2016;96(4):1211–59.

10. Rondina MT, Brewster B, Grissom CK, et al. In vivo platelet activation in critically ill patients with primary 2009 influenza A(H1N1). Chest 2012;141(6):1490–5.

11. Khandaker G, Dierig A, Rashid H, King C, Heron L, Booy R. Systematic review of clinical and epidemiological features of the pandemic influenza A (H1N1) 2009. Influenza Other Respi Viruses 2011;5(3):148–56.

12. Gomez-Casado C, Villaseñor A, Rodriguez-Nogales A, Bueno JL, Barber D, Escribese MM. Understanding Platelets in Infectious and Allergic Lung Diseases. Int J Mol Sci [Internet] 2019;20(7). Available from: http://dx.doi.org/10.3390/ijms20071730

13. Noetzli LJ, French SL, Machlus KR. New Insights Into the Differentiation of Megakaryocytes From Hematopoietic Progenitors. Arterioscler Thromb Vasc Biol 2019;39(7):1288–300.

14. Yang M, Ng MHL, Li CK. Thrombocytopenia in patients with severe acute respiratory syndrome (review). Hematology 2005;10(2):101–5.

